# Inferring the proportion of undetected cholera infections from serological and clinical surveillance in an immunologically naive population

**DOI:** 10.1101/2023.11.01.23297461

**Authors:** Flavio Finger, Joseph Lemaitre, Stanley Juin, Brendan Jackson, Sebastian Funk, Justin Lessler, Eric Mintz, Patrick Dely, Jacques Boncy, Andrew S Azman

## Abstract

Most infections with pandemic Vibrio cholerae are thought to result in subclinical disease and are not captured by surveillance. Previous estimates of the ratio of infections to clinical cases have varied widely (2 to 100). Understanding cholera epidemiology and immunity relies on the ability to translate between numbers of clinical cases and the underlying number of infections in the population. We estimated the infection incidence during the first months of an outbreak in a cholera-naive population using a Bayesian vibriocidal antibody titer decay model combining measurements from a representative serosurvey and clinical surveillance data. 3,880 suspected cases were reported in Grande Saline, Haiti, between 20 October 2010 and 6 April 2011 (clinical attack rate 18.4%). We found that more than 52.6% (95% Credible Interval (CrI) 49.4-55.7) of the population ≥2 years showed serologic evidence of infection, with a lower infection rate among children aged 2-4 years (35.5%; 95%CrI 24.2-51.6) compared with people ≥5 years (53.1%; 95%CrI 49.4-56.4). This estimated infection rate, nearly three times the clinical attack rate, with underdetection mainly seen in those ≥5 years, has likely impacted subsequent outbreak dynamics. Our findings show how seroincidence estimates improve understanding of links between cholera burden, transmission dynamics and immunity.

**Key Results:**

- We combine serological and clinical cholera incidence in an outbreak in a naive population in Grande Saline, Haiti.
- The rate of infection with *Vibrio cholerae* was several times the clinical attack rate.
- Half of the population ≥2 years showed serological evidence of infection.
- For every reported clinical case ≥5 years old, 3.2 (95% CrI 3.0-3.4) people were infected.
- Children 2-4 years old had a lower infection rate, which was not significantly different from the clinical attack rate.

## Introduction

In its global roadmap to eliminate cholera as a public health threat by 2030, the Global Task Force for Cholera Control (GTFCC) considers at least 47 countries as cholera affected [1], where 2.86 million cases (uncertainty range 1.3m to 4.0m) and 95,000 deaths (uncertainty range 21,000-143,000) occur each year [2]. In addition to endemic cholera, small and large-scale cholera outbreaks – as in Yemen, where nearly 2,400 deaths were reported in a large outbreak between 2016 and 2018 – are common [3,4]. However, the true number of pandemic *V. cholerae* infections and cases are greatly obscured by the nature of cholera surveillance systems, care-seeking behavior and the natural history of pandemic *V. cholerae*.

In most cholera-affected countries, surveillance is primarily passive, health facility-based and reliant on a suspected case definition that has low specificity [5,6]. Laboratory confirmation of cholera requires shipping stool samples to central laboratories for culture or PCR, and is typically done only for a subset of cases during outbreaks [7,8]. Rapid Diagnostic Tests (RDT) have become available in recent years but are not routinely used [7]. Biases in estimates of the clinical burden of cholera can result from non-systematic use of diagnostic testing and variability in the proportion of people with symptomatic cholera who seek care outside of facilities under surveillance [6,9,10].

Most cholera infections are thought to be asymptomatic or subclinical. They may not be of clinical relevance but the infected may still transmit the bacteria and acquire immunity, which can have important impacts on the epidemiological dynamics of the current or future outbreaks [11–14]. Importantly, the proportion of the population with full or partial immunity can thus not be determined from clinical surveillance alone but is of crucial importance to gauge the future course of an outbreak or to plan vaccination campaigns.

There is growing interest in complementing clinical surveillance with serological data to provide a more complete picture of infections and burden [15–17]. Recent work has shown that cross-sectional serologic data contain sufficient information to estimate the incidence of recent infections [16,18,19]. This quantity is referred to as *seroincidence*. However, the relationship between medically attended clinical incidence and seroincidence, which is shaped by host, pathogen and surveillance system, remains unclear. Furthermore, most evidence comes from hyperendemic settings like Bangladesh, where exposure to cholera is extremely common. Despite the scientific consensus that the number of asymptomatic or mild infections is many times higher than the number of clinically relevant infections, estimates have varied widely (factor 2 to 100), likely as a result of variable case definitions, individual infection histories, different *V cholerae* strains and host factors (Supplementary Material) [11,13,20–27]. Being able to translate between the number of cases observed at health facilities and the true number of infections in the community, or vice-versa, could help improve the public health utility of serosurveillance data and strengthen our understanding of cholera epidemiology and immunity.

Data collected from a representative serosurvey in Grand Saline, Haiti, between March 22 and April 4, 2011, six months after the start of the first cholera outbreak ever reported there, provide a rare opportunity to gain new insights into the relationship between infections with pandemic *Vibrio cholerae* O1 and clinical disease in a population with no previous exposures to the bacterium [15]. In this study we aim to estimate the true incidence of infections with *V cholerae* O1 and the infection to case ratio during the first 6 months of this unprecedented cholera epidemic by combining information from serological and clinical surveillance.

## Methods

### Data and study population

Serologic data were obtained from a previously published representative serosurvey conducted in the municipality of Grande Saline (population 21,131) between 22 March and 6 April 2011, roughly 6 months after the beginning of the outbreak [15,28]. This study included 2,622 participants aged two years and above from 1,240 households, with 2,527 (93%) participants providing serum samples. Vibriocidal antibody titers were measured using an initial 10-fold dilution followed by serial 2-fold dilutions and expressed as 1:20, 1:40, 1:80, 1:160, …, 1:40960. Reported titer values represent the highest dilution with a ≥50% reduction in turbidity compared with a bacteria-only control, indicating inhibition of bacterial growth. The true titer thus lies in the interval between the indicated and the next higher dilution on the scale (interval censoring). The sampling and laboratory analyses have been described previously in Jackson et al. (2013). We log_2_-transformed the reciprocal titer values (a dilution of 1:20 was transformed to log_2_(20) = 4.32, 1:40 to 5.32, 1:80 to 6.32, …) and assigned 0 to titer values below the limit of detection (<1:20). While the only known *V. cholerae* strain circulating in Haiti before the serosurvey was of serotype Ogawa (Centers for Disease Control and Prevention, 2012), vibriocidal titers to serotypes Inaba and Ogawa were detected. Due to known cross reactivity, we use the maximum measured titer across both serotypes (Figure 1A).

**Figure 1:**
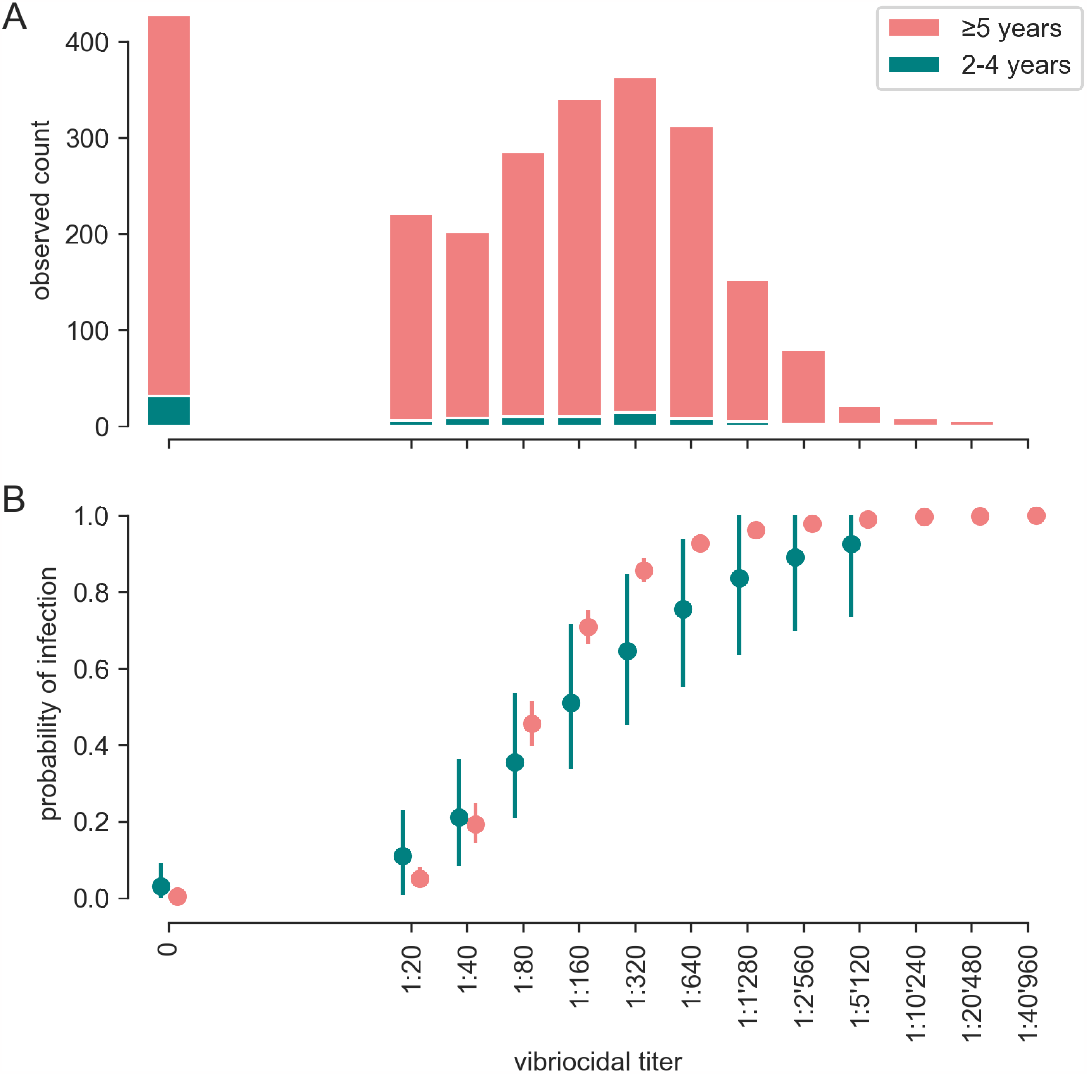
Measured titer values in the sample population (A), inferred individual probability of infection (with 90% prediction intervals) (B) by measured titer value (log_2_-transformed). Note that no estimates of the infection probability for the three highest titer classes for children <5 are shown because no children included in the study had these titers.

In addition to collecting data on individual demographics, study staff asked each participant about episodes of watery diarrhoea occurring since the beginning of the epidemic, if they had sought treatment at a health facility, if they had been hospitalized overnight and if they had been diagnosed with cholera by a health care worker.

We also used the daily incidence of suspected cholera in the commune of Grande Saline reported between 20 October 2010, when the commune reported its first case, and the end of the serosurvey (6 April 2011) from the national surveillance system (Figure 2A) [15,30]. The case definition for suspected cholera used in Haiti at the time was “acute watery diarrhea, with or without vomiting, in persons of all ages residing in an area in which at least one case of *Vibrio cholerae* O1 infection had been confirmed by culture”. The cases were reported in two age groups: children less than 5 years old and individuals aged 5 years and older. In order to compare reported cases to the target population of the serosurvey, which excluded those less than 2 years old, we multiplied the number of reported cases under 5 years by 0.74, the fraction of 2-4 year olds in under 5 year old cases obtained from unpublished age distributions of cases in Haiti in 2010. Our analysis was performed for the entire population 2 years and older and separately for 2-4 year olds and for 5 year olds and older.

**Figure 2:**
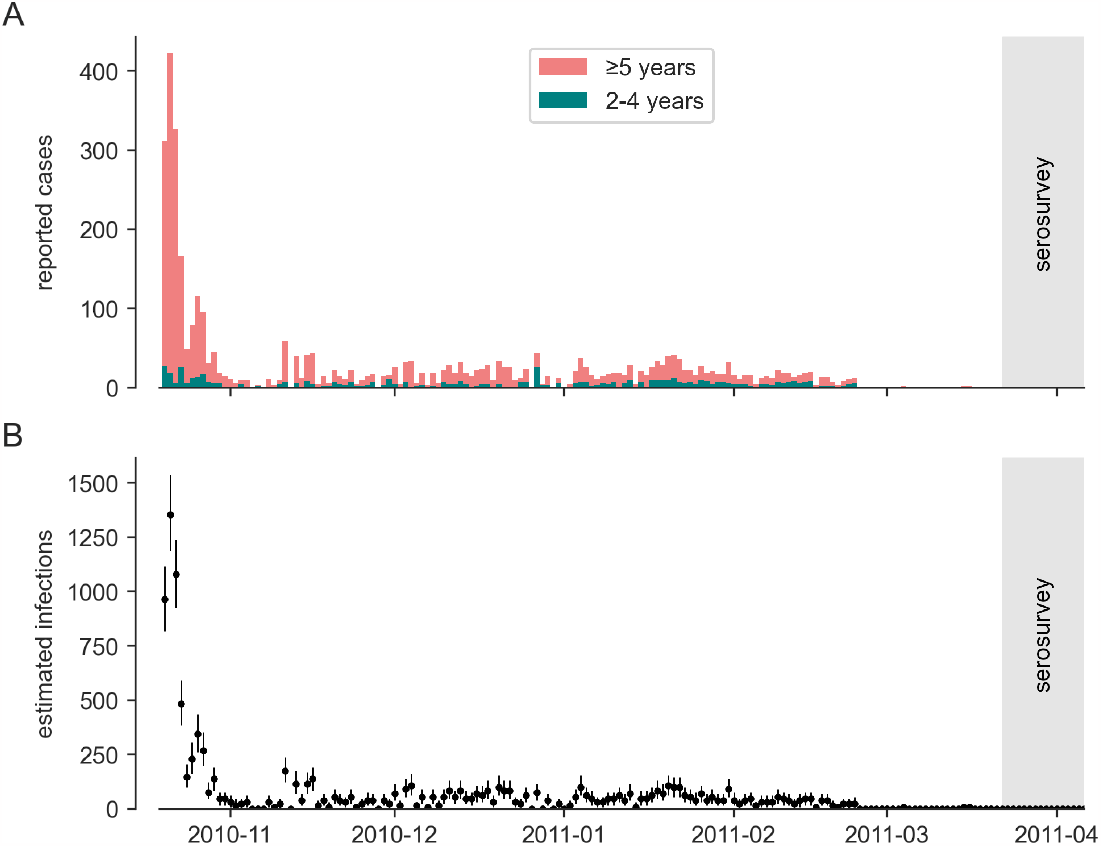
Reported daily clinical incidence of suspected cholera (A) and inferred median incidence of infections with 95% prediction intervals (B).

### Statistical analyses

#### Logistic regression

We evaluated the association of a self-reported cholera diagnosis with vibriocidal antibody titers (log_2_) in the study population using logistic regression, adjusting for age and location. The analysis was performed in R statistical software [31].

### Seroincidence Estimation

#### Bayesian vibriocidal titer decay model

Our primary estimates of seroincidence are based on modeling the vibriocidal titer decay in the blood of participants enrolled in the study. Vibriocidal titer values correlate with recent *V. cholerae* infection and have been shown to be a non-mechanistic correlate of protection when paired samples are available [19,32]. They rise to their maximal value within 1–2 weeks after infection and then start decaying toward pre-infection levels [32]. We developed a hierarchical, Bayesian statistical model of the decay of the vibriocidal titer of participants, from the inferred date of infection to the date the serum sample was taken. As described below, by modeling each individual’s probability of being infected, time of infection, titer decay and titer measurement process, we were able infer the infection rate (i.e., seroincidence) of cholera in Grande Saline from a snapshot of sampled vibriocidal titer values. We assumed that a constant proportion of infections was reported throughout the outbreak, equal incubation periods and equal reporting delays for all individuals, equal infection risk for all individuals (ignoring e.g. household structure or geography) and that infection led to seroconversion in all individuals. For each age group, we fitted the model to the number of individuals at each vibriocidal titer obtained from Jackson et al. (2013).

#### Modeling infection

Our model assumes that the true number of infections each day was proportional to the reported clinical incidence (Figure 2A) lagged by the median incubation period and reporting delay (Table 1). The estimated number of individuals infected on day *t*_*k*_, denoted *I*(*t*_*k*_) is thus:

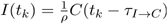

where *C*(*t*_*k*_) is the number of reported cases on day *t*_*k*_ (Figure 2A), *ρ* is the proportion of infections that are reported (a combination of the proportion of infections that are clinically apparent and of the proportion of clinically apparent infections that seek care at reporting facilities) and *τ*_*I→C*_ is the delay from infection to reporting. For each individual *i*, we modeled an infection indicator variable *δ*^*i*^, which takes the value 1 if individual *i* was infected between the beginning of the outbreak and the time of sampling and 0 otherwise, as a Bernoulli process with probability of success equal to the infection rate:

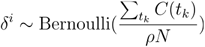

where *N* is the population size.

**Table 1.** Key parameters and priors of the vibriocidal titer decay model.

| Name | Description | Calibrated | Prior / Value |
| --- | --- | --- | --- |
| $N_{pop}$ | population size | No | 21,131 [28] |
| $N_{2-4}$ | population size 2-4 years old | No | $1,588 = (N \cdot 0.1253 \cdot 3/5)$ (estimated from 12.53% under 5s in the 2010 national age distribution and assuming uniform age distribution in the population under 5 [28,39]) |
| $N_{5+}$ | population size, 5 years and older | No | 19,403 (estimated from 2010 national age distribution: 87.47% of the population of over 5 [28,39]) |
| $p_{2-4}$ | proportion of reported cases under 5 that are between 2 and 4 years old | No | 73.94% derived from unpublished age structure of cases in Nov and Dec 2010 |
| $\rho$ | reporting fraction | Yes | Weakly informative prior $\rho \in [\sum_{t_k} C(t_k)/N, 1.5]$ |
| $\sigma$ | titer measurement standard deviation | Yes | $\mathcal{N}_{\geq 0}(0, 1)$ |
| $\bar{\omega}^i$ | Individual baseline titer | Yes | $\begin{bmatrix} \bar{\omega}^i \\ \lambda^i \end{bmatrix} \sim \mathcal{MVN}_{\geq 0} \left( \begin{bmatrix} 5.31 \\ 5.45 \end{bmatrix}, \begin{bmatrix} 3.96 & -4.02 \\ -4.02 & 8.10 \end{bmatrix} \right)$ [19] |
| $\lambda^i$ | Individual titer rise | Yes | |
| $r$ | titer decay rate | Yes | $\mathcal{N}_{\geq 0}(0.0056, 0.0005)$ [$\text{day}^{-1}$]: mean half-life of 123 days [19] |
| $d$ | time from infection to peak titer | No | 11 days [19] |
| $\tau_{I \rightarrow C}$ | time from infection to reporting | No | 2 days: 1.5 days incubation [40] + $\frac{1}{2}$ day of reporting delay |

The time of infection 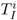 for each infected individual *i* is drawn from a distribution with the shape of the estimated infection incidence from (lagged) reported cases:

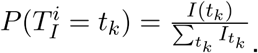

#### Modeling vibriocidal titer dynamics

Following previous studies, we modeled the vibriocidal titer dynamics of infected individuals by assuming an initial increase in titer *d* days after the presumed infection followed by an exponential decay [19,33]. Titers of non-infected individuals were assumed to remain at their baseline level. The modeled *true* titer value of individual *i* at the time of sampling *T*_*s*_ is thus:

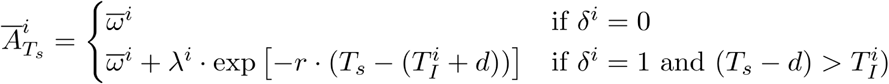

where 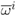 represents the baseline titer value of each individual *i, λ*^*i*^ is the individual temporary titer rise after infection *r* and the exponential decay rate. We do not explicitly model the dynamics of post-infection titer rise, justified by the absence of reported cases in the days immediately preceding the serosurvey (Figure 2A).

#### Measurement model

The measured vibriocidal titer lies on 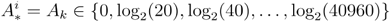 a discrete scale dictated by serial dilutions of the measurement process (see Data). Following Salje et al. (2018) we modeled the probability that the true (log_2_) vibriocidal titer 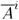 for individual *i* is measured in bin *A*_*k*_ as:

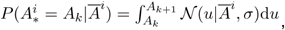

where the width of the log_2_ titer measurement intervals *A*_*k+1*_ − *A*_*k*_ is equal to 3.32 if *k* = 0, corresponding to the initial 10-fold dilution, and 1 otherwise, corresponding to a 2-fold dilution. Measurement error was assumed to be normally distributed on the log-scale with standard deviation *σ*.

#### Parameters

We allowed parameters for titer rise upon infection *λ*^*i*^ and baseline titer 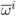 to vary between individuals, whereas the titer decay rate *r* was assumed to be the same for the entire population (Table 1). We used a weakly informative prior for the fraction of reported infections *ρ*, ranging from 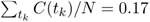 – every inhabitant has been infected – to 1.5, an unlikely over-reporting of 50%. Prior distributions on decay parameters *δ, λ*^*i*^ and 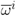, were derived from Jones et al. [19], human challenge studies and cohorts of post-infection cholera cases [16,34]. In addition, we also inferred latent variables representing the infection history of each individual (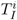 and *δ*^*i*^).

#### Inference

We implemented the model using the PyMC framework for probabilistic programming [35] and calibrated it with the No-U-Turn Sampler (NUTS) [36] with 4 chains of 10’000 tuning iterations and 2’000 posterior sampling iterations. Posterior predictive checks and diagnostic checks are presented in the Supplementary Material. We ran separate inference for the entire population and for the different age groups and report mean and 95% credible intervals (CrI) of the resulting distributions.

#### Ethics and Reproducibility

Participants in the original study provided written informed consent and the study protocol was approved by the Haitian National Ethics Committee IRB (Protocol #2011-FWD-CHOLERA-01) and the U.S. Centers for Disease Control and Prevention IRB (CDC Protocol #6038). This secondary analysis was approved by the Ministère de la Santé Publique et de la Population de l’Haïti and deemed to be exempt from human subjects research by the Johns Hopkins Bloomberg School of Public Health IRB.

The code and data are available on Github (github.com/HopkinsIDD/grande-saline-cholera-serosurvey) and Zenodo.

## Results

In the commune of Grande Saline a total of 3,880 suspected cholera cases (clinical attack rate 18.4%) were reported from the first reported case on 20 October 2020 to the end of the serosurvey on 6 April 2011 through the national cholera surveillance system. Age specific clinical attack rates were 16.4% in people 5 years and above 39.5% in children under 5. We estimated the clinical attack rate to be 39.5% in children 2-4 years old (Table 2).

**Table 2:**
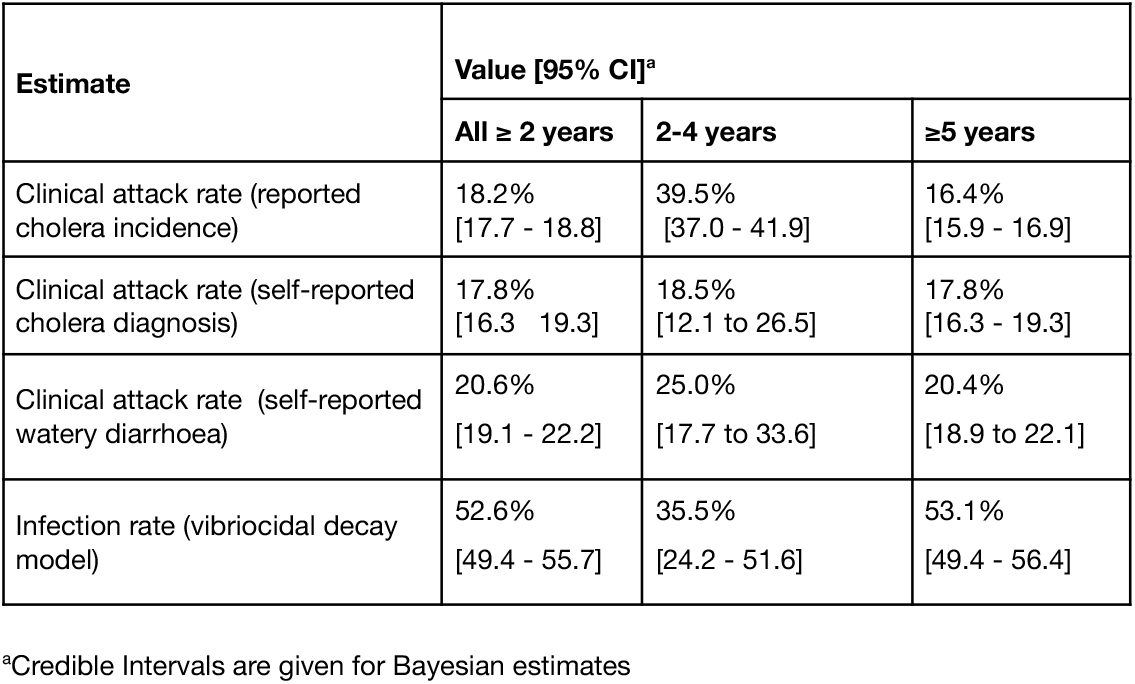
Clinical attack rate, attack rate of self reported cholera and watery diarrhoea, and estimates of infection rate with *V. cholerae* O1 according to our main and alternative analyses.

| Estimate | Value [95% CI]<sup>a</sup> |  |  |
| --- | --- | --- | --- |
|  | All ≥ 2 years | 2-4 years | ≥5 years |
| Clinical attack rate (reported cholera incidence) | 18.2%<br>[17.7 - 18.8] | 39.5%<br>[37.0 - 41.9] | 16.4%<br>[15.9 - 16.9] |
| Clinical attack rate (self-reported cholera diagnosis) | 17.8%<br>[16.3 - 19.3] | 18.5%<br>[12.1 to 26.5] | 17.8%<br>[16.3 - 19.3] |
| Clinical attack rate (self-reported watery diarrhoea) | 20.6%<br>[19.1 - 22.2] | 25.0%<br>[17.7 to 33.6] | 20.4%<br>[18.9 to 22.1] |
| Infection rate (vibriocidal decay model) | 52.6%<br>[49.4 - 55.7] | 35.5%<br>[24.2 - 51.6] | 53.1%<br>[49.4 - 56.4] |

<sup>a</sup>Credible Intervals are given for Bayesian estimates

Five months after the peak of reported cases in the first wave, 2,622 people 2 years and older were enrolled in a representative serosurvey [15], 4.7% (124) were 2-4 years of age and 59% (1,553) were female. 20.6% (541) of participants reported watery diarrhoea since the start of the outbreak in October 2010, and 17.8% (466) reported to have been diagnosed with cholera by a health care worker. 14.8% (388) of participants reported to have sought treatment at a health facility with half of them (50.5%, 196/388) having spent at least one night there (Table 2). Having a self-reported cholera diagnosis or watery diarrhoea were both independently associated with higher vibriocidal log_2_-titer levels in multivariable analyses (odds ratios 1.12 (95% CI 1.08 to 1.16) and 1.10 (95% CI 1.07 to 1.14), Supplementary Material). Among the 2,526 participants with reported vibriocidal titer values, 18.2% had no detectable vibriocidal antibodies and 38.8% had a titer of ≥320 to either of the two serotypes.

We combined vibriocidal titers and reported clinical incidence to infer the infection to reported case ratio and the infection rate. We found a sigmoidal dose-response relationship between titer and the probability of having been infected. People with a titer of 40 had a probability of 18.5% (95% CrI 13.2-24.1) of having been infected and those with a titer of 320 had a 85.8% (95% CrI 82.6-88.9) probability. When stratifying by age, we found that the curves had similar shapes between children 2-4 years old and those 5 and older (Figure 1B).

We estimate that 52.6% (95%CrI 49.4-55.7) of Grande Saline’s population ≥2 years old was infected during the first 6 months of the cholera epidemic, including 35.5% (95%CrI 24.2-51.6) of children 2-4 years old and 53.1% (95%CrI 50.1-56.4) of those aged 5 and above (Table 2, Figure 2B and Figure 3A). When comparing these estimates to the incidence of reported clinical cholera cases, we find that the implied infection to reported case ratio was far higher in older children and adults than among those 2-4 years old. On average we estimate that for every reported clinical case among those ≥5 years old, a total of 3.2 (95% CrI 3.0-3.4) people were infected. In contrast, for every reported clinical case among those 2-4 years old there were 0.9 (95% CrI 0.6-1.3) infections (Figure 3B).

**Figure 3:**
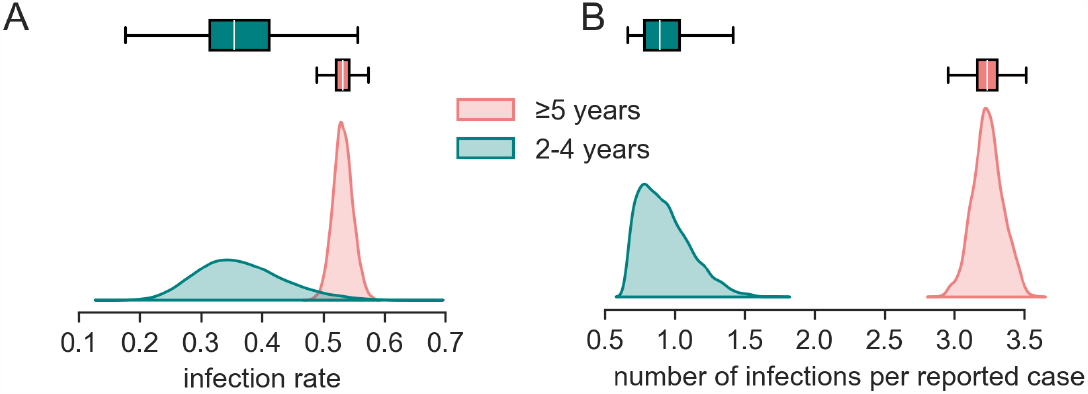
Distribution of the infection (symptomatic and asymptomatic) to reported case ratio (A) and the infection rate (B) in Grande Saline during the study period inferred from combining serological and incidence data and modeling vibriocidal decay.

## Discussion

We estimated that more than 52.6% (95% CrI 49.4 to 55.7) of the population in Grand Saline, Haiti was infected by *V. cholerae* O1 during the first 6 months of the outbreak in 2010, with young children (2-4 years old) having a 32.5% (95% CrI 2.4 to 54.1) lower seroincidence than those 5 years and older, despite both age groups being immunologically naive at the start of the outbreak. Compared to the reported clinical incidence from the same community, we found that for every three infections among those five years and older in the community one clinical case was reported through the national surveillance system, though this ratio was closer to one for those 2-4 years old. As the global community looks for new ways to understand the burden and transmission of cholera, these estimates provide a unique perspective to help interpret clinical and seroincidence especially in immunologically naive populations.

Our estimates of the ratio of infections to reported clinical cholera cases are at the lower end of those previously reported, which likely reflects that this population had no previous exposure to the bacteria, but could also reflect differences in host factors (e.g., undernutrition) and exposure routes [11–13,20,21,24–27]. Our estimated number of infections among young children nearly matches the numbers of reported suspected cholera cases, this either implies that there were almost no infections in this age group that did not results in medically-attended disease, or more realistically that unreported infections got compensated by the poor specificity of the suspected cholera case definition, especially in young children, where other etiologies frequently lead to similar symptoms.

Given that other diarrheal pathogens may also have distinct seasonality and patterns of immunity (e.g., rotavirus [37]), our infection to reported case ratio may not be as generalizable across time and geographies as the estimate for adults.

Our study comes with a number of limitations. In reconstructing the infection incidence, our estimates of vibriocidal boost and decay among all infected individuals, across the spectrum of clinical severity and age, were based on priors informed from estimates from a cohort of severe, mostly adult, cholera cases in Bangladesh. While to our knowledge there are no detailed kinetic data available across age and severity, especially among immunologically naive, it may be that mild infections lead to a more blunted immune response, as seen with other pathogens [38], and that young children have different boosts and decay rates from adults. In estimating infection incidence we only relied on vibriocidal titers, with no other markers considered. Recent work has shown that inclusion of multiple serologic markers (i.e. antibodies against cholera toxin) can improve estimates of seroincidence from cross-sectional data [16,19], and inclusion of these in a multivariate decay model could have improved our precision. However, we decided not to include data on anti-cholera toxin B antibodies as they were measured using an assay for which no post-infection dataset to parametrize the kinetic model were available, in addition to the computational complexity of simultaneously modeling the decay of multiple antibodies. We assumed that the hazard of infection was proportional to the observed epidemic curve of suspected cholera cases, and that this ratio was constant in time. Although the agreement of attack rate estimates from clinical surveillance and self-reported cholera indicates that surveillance performed well, the surveillance system may have changed over the course of this six month period given the unprecedented context, and people’s care seeking behaviors may have changed as awareness of cholera increased over the course of the epidemic.

Our estimates of the number of infections per reported suspected cholera case during an outbreak in an immunologically naive population suggests that only one in three infections with pandemic *V cholerae* resulted in medically attended clinical disease that was reported via the surveillance system. More than half of the population of Grande Saline thus likely acquired at least partial immunity to cholera with unknown duration, a result which, given the similar reported clinical attack rates and similar surveillance system, can likely be generalized to large parts of Haiti. This has likely impacted the dynamics of the 2011 and subsequent epidemic waves. Although estimates may not be generalizable to populations with more frequent historical exposures to *V cholerae*, this study shows that population immunity to cholera cannot be understood based on clinical surveillance data alone. While more work is needed to grasp the links between infections, immunity and its duration across different populations, our results highlight the potential utility of combining clinical and serologic data in assessing cholera risk and making resource allocation decisions for cholera prevention and control.

## Acknowledgements

The findings and conclusions of this report are those of the authors and do not necessarily represent the official position of the Centers for Disease Control (CDC). FF started to work on this analysis when at the London School of Hygiene and Tropical Medicine. He currently works for Epicentre.

