## Supplementary Material for "Inferring the proportion of undetected cholera infections from serological and clinical surveillance in an immunologically naive population"

### Previous estimates of infection to clinical case ratio

Table S1: previous published estimates of the cholera infection to clinical case ratio

| **Location and year** | **Study design & population** | **Method to determine infection** | **Biotype** | **Estimate** | **Reference** |
|---|---|---|---|---|---|
| East Pakistan, 1966–1967 | Longitudinal study of all available village residents in an endemic area | Serology | El Tor | 0 out of 27 infected required treatment<br><br>22 out of 27 infections inapparent | (Mccormack, Islam, and Fahimuddin 1969) |
| East Pakistan, 1968-1969 | Case contact follow-up | Bacterial culture & serology | Classical | Infection to case ratio 4:1 | (Bart et al. 1970) |
| | | | El Tor | Infection to case ratio 36:1 | |
| (unspecified) | Surveys | Bacterial culture | Classical | 59% inapparent infections and 15% very mild symptoms but detected only in bacteriological surveys | (Gangarosa 1974) |
| | | | El Tor | 75% inapparent infections and 18% very mild symptoms but detected only in bacteriological surveys | |
| Louisiana, US, 1978 | Case contact follow-up | Bacterial culture | El Tor | 3 out of 6 infections inapparent | (Blake et al. 1980) |

| Location and year | Study design & population | Method to determine infection | Biotype | Estimate | Reference |
|---|---|---|---|---|---|
| Review of published sources | Case contact follow-up | Bacterial culture | Classical | 50% of infected were asymptomatic (mean over 5 studies)[1] | (Feachem 1982) |
| | | | El Tor | 70% of infected were asymptomatic (mean over 4 studies)[1] | |
| Gulf Coast Oil Rig, US, 1983 | Outbreak investigation | Serology | El Tor | 1/16 infections inapparent | (Johnston et al. 1983) |
| Truk, Micronesia, 1982 | Cross-sectional study | Serology | El Tor | 68% of infections inapparent | (Harris et al. 1986) |
| Review of published sources | - | - | (not specified) | 1/2 to 1/100 infections leads to symptomatic cholera in endemic areas | (Glass and Black 1992) |
| Bay of Bengal, 1891–1940 | Model of historical incidence of clinical cholera | - | Classical | high proportion of inapparent infections | (King et al. 2008) |
| Sitakunda, Bangladesh, 2022 | Longitudinal study | Serology | El Tor | 1 symptomatic case per 600 infections<br><br>1 medically-attended case per 2,340 infections | (Hegde et al. 2023) |

[1] The studies included in this aggregated estimate are not mentioned separately in this table.

### Predictors of self-reported cholera diagnosis

Using multivariate logistic regression and adjusting for age, we found that self-reported cholera diagnosis in the participants of the serosurvey was associated with a higher $log_2$-transformed vibriocidal Ogawa titer (Odds Ratio (OR) 1.10 (95% CI 1.06 to 1.13), p-value <0.001), higher values of IgG to heat labile toxin (OR 3.26 (95% CI 2.22 to 4.80), p-value <0.001) and a higher ratio of IgG against cholera toxin to IgG against heat labile toxin (OR 1.24 (95% CI 1.13 to 1.38), p-value <0.001). Other covariates (IgA to cholera toxin and heat labile toxin, sex, pregnancy, village) were not included in the final model.

### Alternative methods to infer infection rate

We took two additional approaches to estimating the infection rate to compare to our main model; use of a fixed vibriocidal threshold and Gaussian mixture models.

#### *Fixed vibriocidal antibody titer thresholds to determine recent infection*

We used a previously established vibriocidal antibody titer threshold of 320 to identify people infected with cholera in the previous 200 days (Azman et al. 2019). We accounted for the imperfect sensitivity and specificity of this threshold using the Rogan-Gladen estimator, considering estimates from two different populations, a study among previously uninfected adult North American volunteers challenged with *Vibrio cholerae* O1 and confirmed cholera cases in Bangladesh (Rogan and Gladen 1978). Uncertainty around sensitivity and specificity were propagated through the estimator.

From this approach we estimate that 42.0% (using North American cohort) to 31.2% (95%CI 14.7 to 46.4, Bangladesh cohort) of the population ≥2 years old was infected. While the estimates of the infection rate for 2-4 year olds were slightly lower than those from our main analyses, those for ≥5 were on average 1.6 times lower (Table S2, Figure S1).

#### *Gaussian mixture model to determine recent infection*

We also fit a mixture model composed of two normal distributions to the bi-modal distribution of measured $log_2$-transformed vibriocidal antibody titers of participants. The two normal distributions were interpreted as the baseline antibody titer levels of uninfected and increased titer levels of recently infected participants respectively. We accounted for interval censoring of the titer values and estimated mean and variance of each distribution in addition to the mixing parameter, which we interpreted as the infection rate. Priors for the two normal distributions were Normal(4.5, 5) and Normal(8.5, 5). The prior for the mixture parameter was set to a Beta(1,1) distribution. The model was implemented in Stan using the RStan package (Stan Development Team 2020).

This approach produced similar central estimates than the threshold model for the population aged 5 and above (31.8%; 95% CrI 24.1 to 41.3) and a flat distribution with wide credible interval for 2 to 4 year olds (65.2%; 95% CrI 11.8 to 95.0) (Table S2, Figure S1).

Our supplementary analysis found that our approach to estimating the infection rate led to generally larger estimates than the alternative approaches including the use of a fixed threshold and a mixture model. While commonly used and computationally simpler, neither accounts for the waning of antibody levels in the months post infection. In the absence of well characterized post-infection kinetics or slowly waning antibodies, these approaches may perform well but care should be taken when applying these generally with fast waning antibodies, like vibriocidals (half life ~120 days, (Jones et al. 2022)).

Table S2: Clinical attack rate, attack rate of self reported cholera and watery diarrhoea, and estimates of infection rate with *V. cholerae* O1 according to our main and alternative analyses.

| **Method** | **Value [95% CI or CrI][a]** | | |
|---|---|---|---|
| | **All ≥ 2 years** | **2-4 years** | **≥5 years** |
| **Clinical attack rate estimates** | | | |
| **Clinical attack rate (reported cholera incidence)** | **18.2%** | **39.5%** | **16.4%** |
| Clinical attack rate (self-reported cholera diagnosis) | 17.8% | 18.5% | 17.8% |
| Clinical attack rate (self-reported watery diarrhoea) | 20.6% | 25.0% | 20.4% |
| **Infection rate** | | | |
| **Infection rate computed using a vibriocidal decay model** | **52.6% [49.4 - 55.7]** | **35.5% [24.2 - 51.6]** | **53.1% [49.4 - 56.4]** |
| **Estimate of the infection rate from alternative analyses** | | | |
| Infection rate computed using a fixed vibriocidal antibody threshold and correcting for sensitivity and specificity computed from a cohort from Bangladesh | 31.2% [14.7 - 46.4] | 21% [4.1 - 36.2] | 31.7% [15.1 - 46.9] |
| Infection rate computed using a fixed vibriocidal antibody threshold and correcting for sensitivity and specificity computed from a cohort of healthy North-Americans | 42.0%[b] | 33.9%[b] | 42.3%[b] |

| | | | |
|---|---|---|---|
| Infection rate computed using a gaussian mixture model | 32.0% [24.5 - 41.3] | 65.2% [11.8 - 95.0] | 31.8% [24.1 - 41.3] |

[a]Credible Intervals are given for Bayesian estimates

[b]Uncertainty cannot be quantified because no confidence intervals on sensitivity and specificity of thresholds are given in the source used

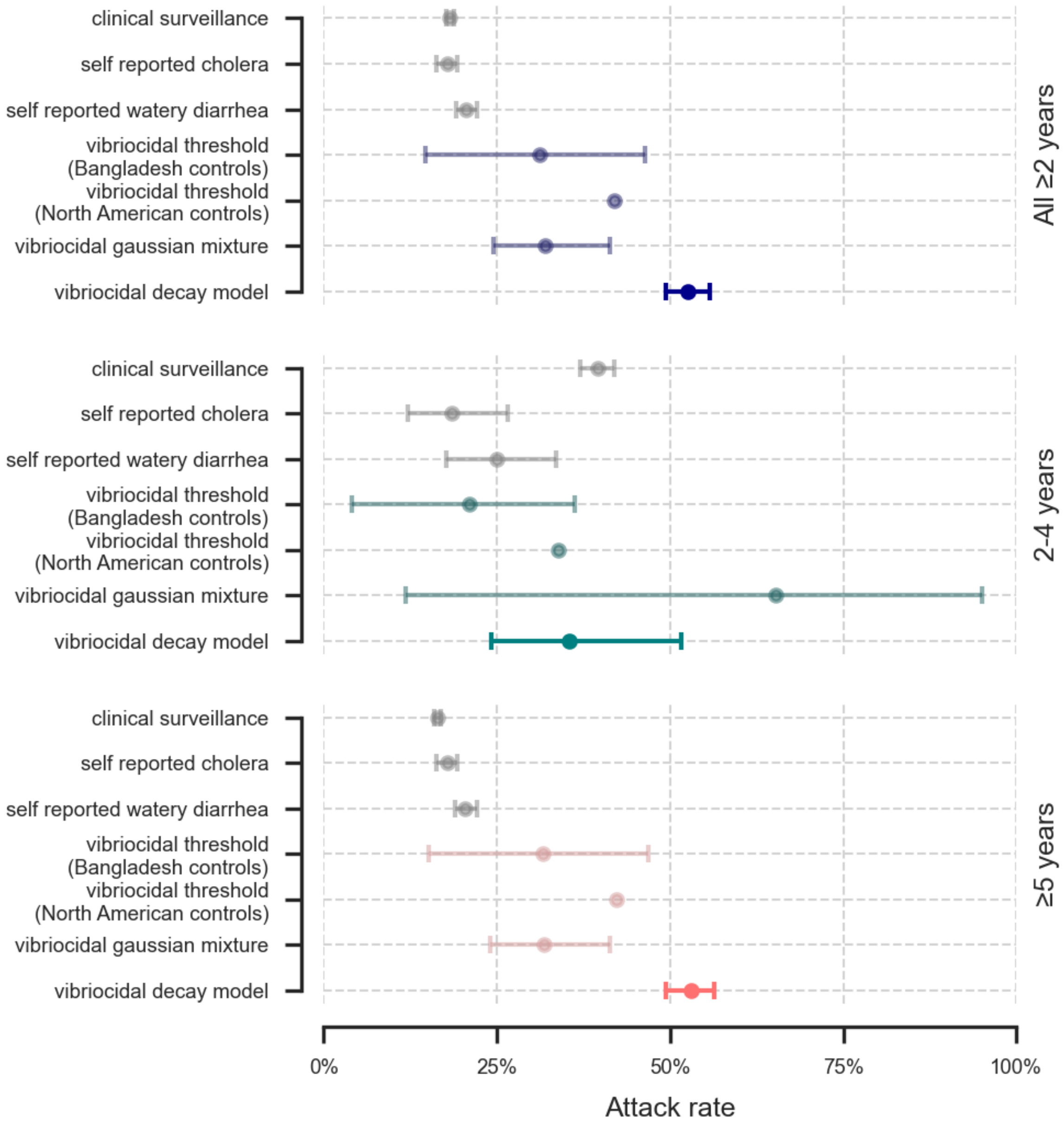


Figure S1: Estimates of clinical attack rate (grey) and infection rates (colored) for age groups 2 years and above, 2-4 years and 5 years and above.

### Bayesian vibriocidal titer decay model

#### *Parameter values and fit*

The vibriocidal titer decay, parametrized by the individual baseline titer $\varpi^i$, the individual titer rise $\lambda^i$ and the titer decay rate $r$, didn't change significantly during the model inference, and the posterior values are similar to the priors from Jones et al. (2022). Samples from the posterior distribution are shown in Figure S2. There is a significant overlap between baseline titer of the uninfected population and titer values of the infected population, even when infected titer values are at their maximum. This overlap explains the sigmoid curve and a part of the uncertainty on our parameter identification.

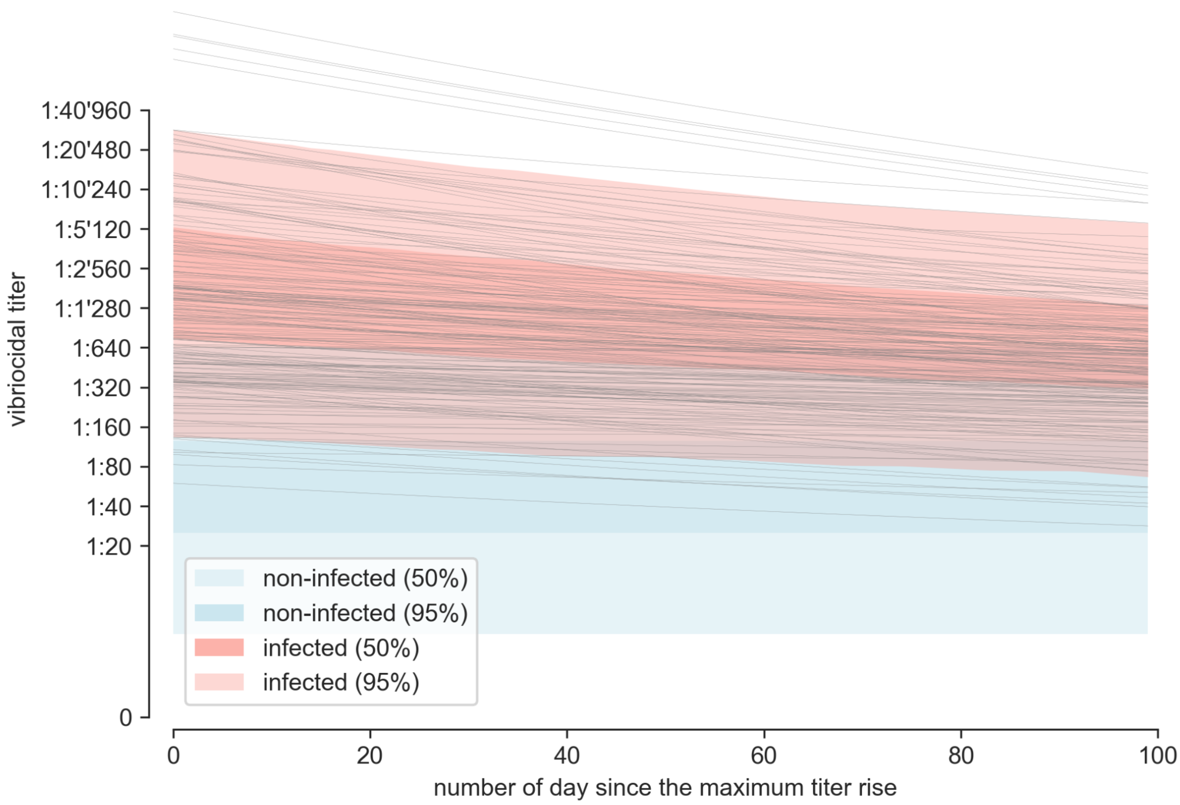


Figure S2: Visualization of the posterior of the vibriocidal titer decay model. The decay of infected individuals (red shade, with 100 samples in gray lines) is shown from the maximum titer rise, 11 days after infection. Uninfected individual titer values remain constant at their baseline level (blue shade).

#### *Posterior predictive checks*

Posterior predictive checks compare the observed data with replicated data from the fitted decay model. The discrepancies between posterior predictive checks and posterior values may come from a inadequacy in the model definition (Figure S3). All observations fall within the range of posterior predictive values, but for the lower titer values, the model underestimates the median count. One possible explanation would be that the discrete latent parameters indicating infection are not inferred correctly, a common pitfall in

Bayesian inference. However, given that the posterior of the Bernoulli draws of the infection indicator $\delta^i$ are correctly distributed with respect to its inferred parameter value, the infection rate, $\mathrm{iAR}$, we ruled out this possibility. It is thus most likely that this bias is due to our model specification and the difficulty to reconcile observed data with the provided decay parameters and the case time series.

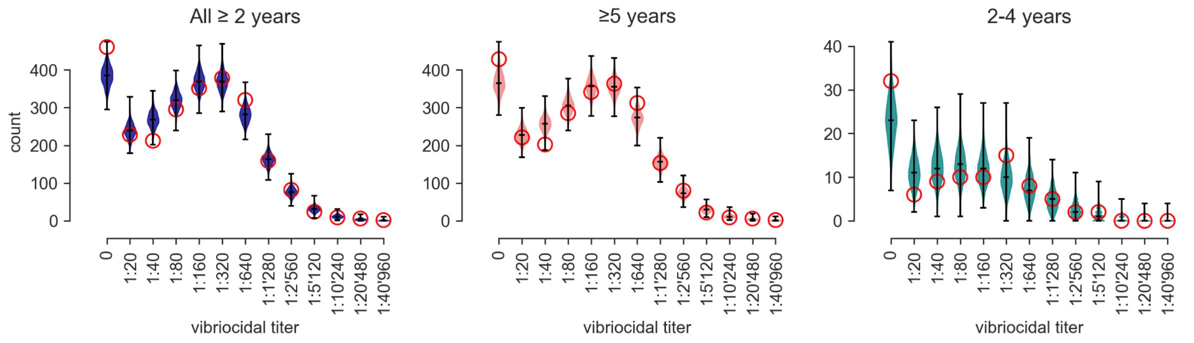


Figure S3: Posterior predictive checks of the three bayesian decay models developed for each age group. The red circle represents the observed count in each titer bin, and the violin plots the posterior predictive values.

#### *Sensitivity analysis on the decay rates of antibody*

The antibody decay rate parameter $r$ is derived from prior studies and informed by a single narrow prior common to all age groups (Table 1). A hypothetical explanation for the observed difference in infection rate and infection-to-reported-case ratio between age groups could be a true decay rate varying by age. We perform a sensitivity analysis, assessing whether a different decay rate in children could explain the differences in infection rate between age groups.

We fit our model for 2-4 years old with a prior on the decay rate multiplied by 1/10, 1/2, 5, and 9. As shown in Figure S4, the differences in decay rate influence our estimates, changing the density and modes of the infection to the reported case ratio and of the infection rate. For example, the mean of the infection to the reported case ratio varied from 0.8 (95%CrI 0.7-1.1) with a multiplier of 1/10 to 1.3 (95%CrI 0.7-2.2) with a multiplier of 9. This variation, however, remains small compared to the variation between age groups. The distinction between the two age groups remains clear even assuming a much faster or slower decay rate for children. The different infection rates and infection to reported case ratios for the two age groups are thus not an artifact of our choice of the decay rate prior.

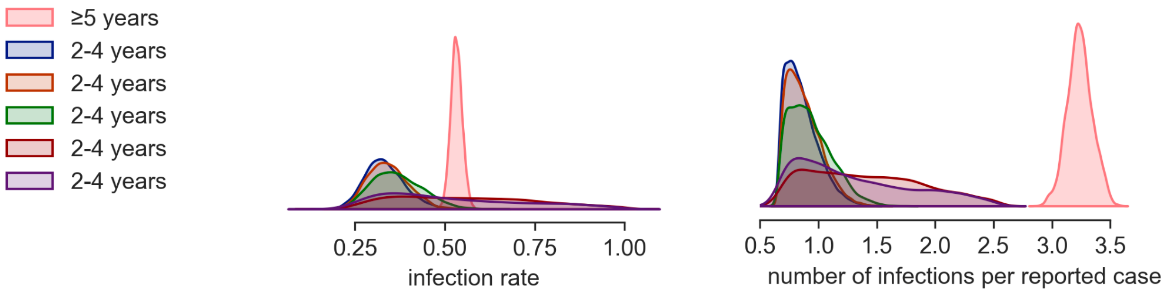


Fig S4: infection to reported case ratio and infection attack date derived from our main model for 5 and older, and from a model with different priors on the decay rate parameter for the individuals 2-4 years old.
